# Modelling The Spread of COVID-19 Using The Fundamental Principles of Fluid Dynamics

**DOI:** 10.1101/2020.06.24.20139071

**Authors:** Harris Sajjad Rabbani, Kofi Osei-Bonsu, Jassem Abbasi, Peter Kwame Osei-Bonsu, Thomas Daniel Seers

## Abstract

The increasing number of positive cases of SARS-CoV-2 worldwide led to a catastrophic breakdown in the global economy and unprecedented social disruptions. These measures have significantly impacted the world’s economy and in many cases has led to the loss of livelihood. Mathematical modeling of pandemics is of critical importance to understand the unfolding of transmission events and to formulate appropriate control measures. In this article, we introduce a physics based approach for forecasting epidemics such as COVID-19. The proposed physics-based mathematical model stems from the fundamental principles of fluid dynamics, and can be utilized to make projections of the number of infected people at any scale. Our model takes into account the diffusive transmission of the virus, the growth of virus inside the human body and the response of the natural immune system of individuals. We demonstrate that the health of individuals plays a critical role in controlling the evolution of the epidemic. In places where the individuals exhibit a strong immune system, the development of pandemic is limited despite high diffusivity of the virus. Overall, the proposed mathematical model can be beneficial for predicting and designing potential strategies to mitigate the spread and impact of pandemics.

## 1 Introduction

In December 2019, a cluster of cases of pneumonia, subsequently associated with a novel coronavirus (Severe Acute Respiratory Syndrome - Coronavirus-2, SARS-CoV-2), named Coronavirus Disease 2019 (COVID-19) by the World Health Organization (WHO), emerged in Wuhan, China. It was rapidly declared a pandemic on March 11, 2020, in view of its exponential spread worldwide ^[1]^. As of 23^th^ October 2020, the WHO had reported 41.9 million confirmed cases and over 1.16 million deaths globally, with the highest number of cases reported in the United States of America (USA) ^[2]^. The rapid spread of the virus continues to pose a monumental global health challenge.

Clinically, infected subjects exhibit a wide range of non-specific features, from mild-to-moderate symptoms such as cough, fever and fatigue to severe, life-threatening respiratory and systemic complications. On the other hand, it has been well documented that infected persons may exhibit no symptoms at all (asymptomatic) or may be yet to manifest symptoms (pre-symptomatic), but still potentially infectious ^[3-4]^. In such cases, infected individuals may be likely to maintain normal social interactions, without realizing the need for self-isolation due to the obscurity of their symptoms.

Currently, our understanding of the transmission risk is incomplete. Epidemiologic examination in Wuhan at the beginning of the outbreak identified an initial connection with a live animal seafood market, where patients had worked or visited ^[5]^. As time progressed, person-to-person spread became the main mode of transmission ^[6]^. Although SARS-COV-2 has been detected in non-respiratory samples such as stool and blood, the transmission is primarily thought to occur through close contact, via respiratory droplets and aerosols ^[7]^. The virus released in these secretions when an infected subject coughs, sneezes, or talks can then infect another person if it makes direct contact with the mucous membranes ^[8-9]^.

Under ideal circumstances, an effective vaccine might be administered to mitigate the dire effects of the virus. Regrettably, the development of an acceptable vaccine to this end appears unlikely in the short-term. Consequently, government and public health responses have focused mainly upon non-pharmacological interventions ^[10]^. These measures include physical/social distancing to minimize the rate of person-to-person contact, frequent hand washing, the utilization of masks, gloves and other forms of personal protection equipment (PPE), mass testing, contact tracing and isolation/quarantine of persons with suspected and confirmed cases of COVID-19 infection.

Although these interventions have contributed significantly to the gradual decline of the transmission rate and by extension deaths worldwide, there are increasing concerns that the easing of these measures may result in the surge of new cases ^[10]^.

In pandemic situations where data could be sparse, mathematical modelling can be a powerful tool to understand and predict the course of the outbreak in order to inform the development of potential control strategies ^[11-12]^. The most frequently used framework in the case of human transmissions is the so-called SIR model ^[13]^. According to this model, the individuals are categorized into three groups: susceptible *S*, infected *I* and recovered *R*. Mathematically, the transition of individuals among these three groups is computed using the Ordinary Differential Equation (ODE) to predict the overall behavior of the number of infected persons. Several, more complex variants of the SIR model have been developed in an attempt to capture the transmission dynamics of pandemics more accurately. Particularly in the case of COVID-19, several modifications have been made to the SIR framework to consider the number of deceased ^[14]^ and the effect of public health containment policies ^[15]^, including the fraction of undocumented infections and their contagiousness ^[16]^. Wu et al. ^[17]^ utilized available data to model the case fatality risks of symptomatic persons in Wuhan, China. Giordano et al. ^[18]^ further extended the model to distinguish between detected and undetected cases and the level of severity of the manifested symptoms during the course of the outbreak.

In this article, we introduce an infectious modeling technique to forecast disease outbreaks, specifically COVID-19, using the fundamental principles of fluid dynamics. Simplistically, we consider a carrier of the virus as a fluid containing a dissolved ionic species. With this intuition, we attempt to derive a simplified theoretical model using the well-known Fluid Transport Equation ^[19]^ to predict the transmission and propagation of COVID-19. The proposed physics based mathematical model takes into account the diffusive transmission of the virus, the growth of virus inside the human body and the response of the natural immune system of individuals. Our analysis demonstrates that a strong immune system could be critical for mitigating the impact of the pandemic. To our knowledge, a physics based model of this kind has not been discussed in the literature.

## 2 Results and Discussion

Using the fluid transport equation, we derived the following mathematical model to predict the infected cases of COVID-19. The details concerning the derivation of mathematical model are provided in the methods section. 

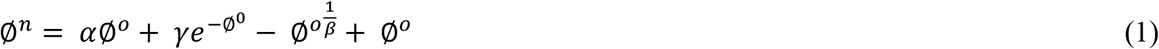

The number of infected cases is given by Equation 1 where ∅ is the ratio of total infected cases *I*, over the initial number of susceptible people *S*^*o*^(assumed as the entire population of the area). There are three dimensionless fitting parameters in Equation 1 *α, γ* and *β. α* is a diffusion factor that takes into account the diffusive transmission of infection due to person-to-person contact; *γ* describes the growth of the virus within an individual and *β* relates to the immune system of the individual. The subscripts *n* and *o* refer to the new and old cases respectively. The detailed description of fitting parameters used to solve the model (Equation 1) is given in Table 1. The visual representation of the fitting parameters is shown in Figure 1.

**Table 1.** Definition of fitting parameters that are used to solve the presented physics based model.

| Parameters | Physical Description |
| --- | --- |
| $\alpha$ | The diffusive transmission of the virus from person to person. |
| $\gamma$ | The growth of the virus within an individual. |
| $\beta$ | Describes the effectiveness of the immune system of the human body to resist the growth of the virus. |

**Figure 1.**
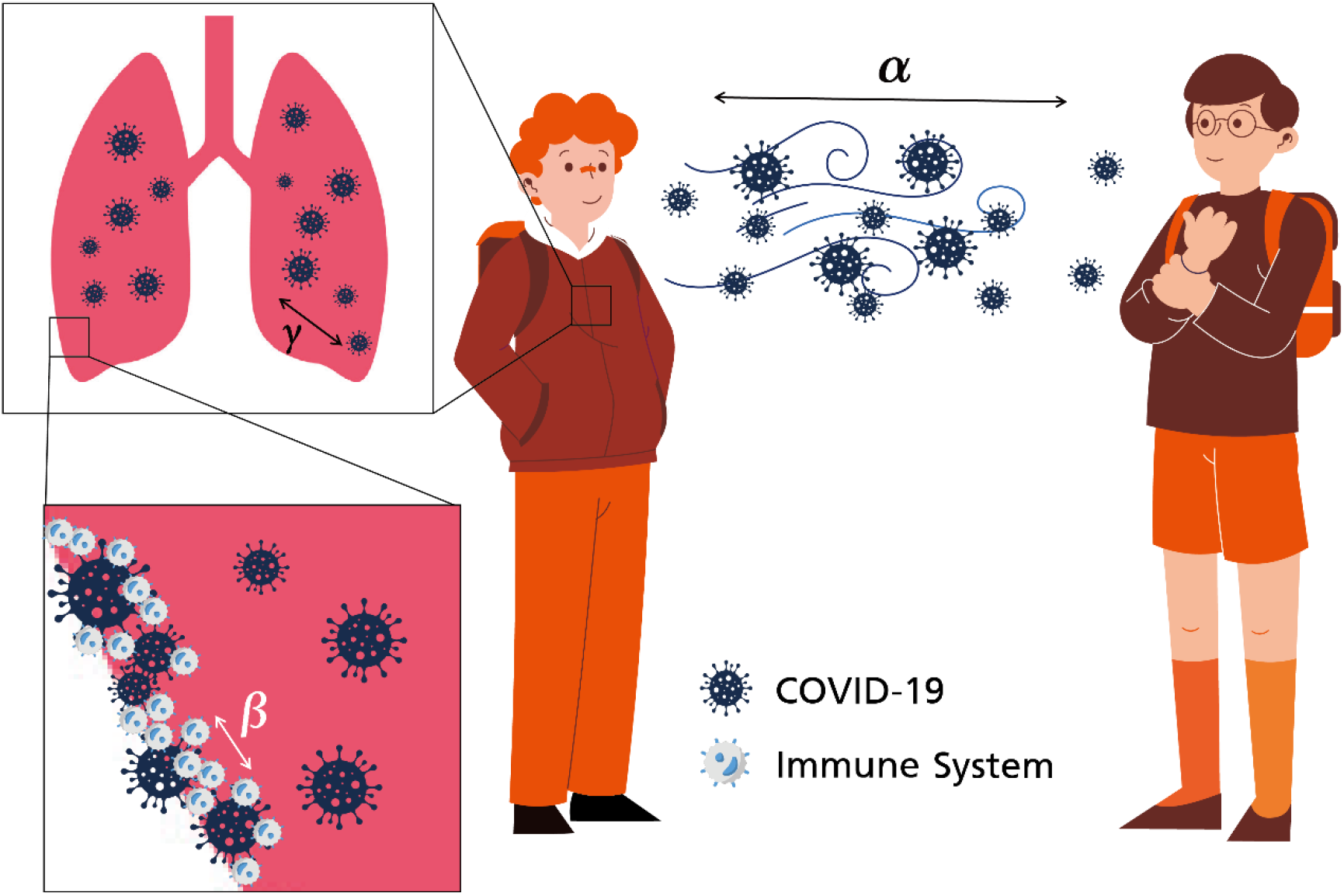
Cartoon depicting the physical meaning of the fitting parameters represented in the model. *α* relates to the diffusive transmission of the virus, *γ* is the activity of virus within the individual and *β* is related to the strength of the individual’s immune system. Both the environmental conditions as well as human interventions such as social distancing and wearing of protective gears (e.g. masks) can have influence *α*. Whereas the medical condition of individuals affects the *γ* (the growth of virus) and the *β* (natural immunity) values.

The value of ∅ for each country as of 27^th^ July 2020 provided by the EU Open Data Portal ^[20]^ is shown on the world map (Figure 2 (a)). It can be seen from Figure 2 (a) that the northern and the western part of the world is heavily affected by COVID-19, whereas in Oceania, Asia and Africa, the pandemic is relatively less severe. Figure 2 (a) suggests that the environmental changes across the continents can play a significant role in the spread of COVID-19 ^[21]^. Figure 2 (b) shows the comparison of the trend of infected cases as computed by the model with recorded data for 16 countries provided by EU Open Data Portal ^[20]^ as of 27^th^ July 2020. Overall, the model (Equation 1) is able to capture the trend of the cases and also forecast future cases. The sensitivity analysis of fitting parameters is illustrated in Figure 2 (c-e). Increase in *β* reduces the number of infected cases, whereas an increase in *α* and *γ* increases the number of infected cases.

**Figure 2.**
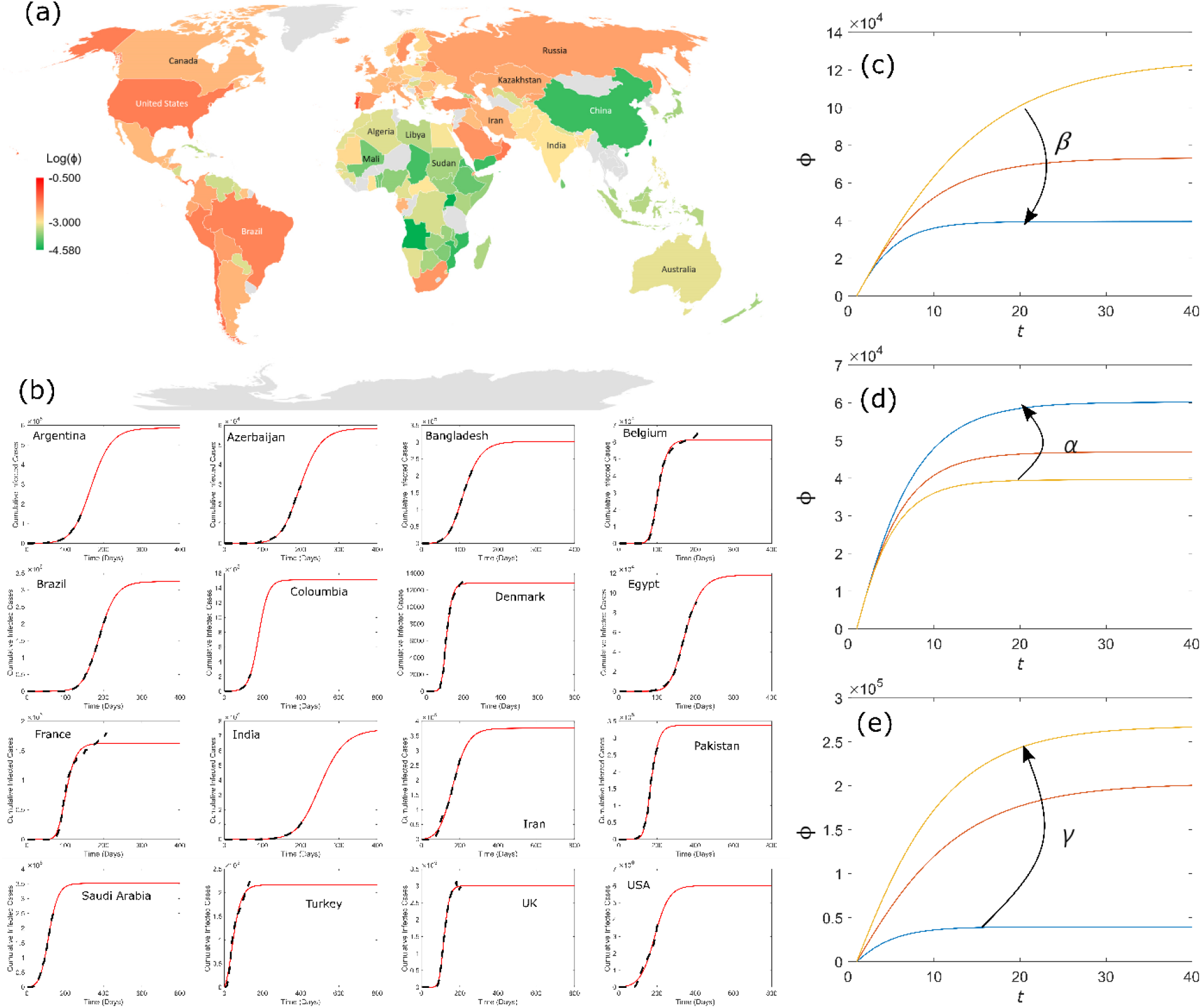
(a) Worldwide distribution of COVID-19 as of 27^th^ July 2020 provided by the EU Open Data Portal (20). In Asian and African countries the epidemic is less severe than American and European countries. This localized severity of COVID-19 in the northern and western part of the globe may reflect the role of the environment on the impact of the pandemic. (b) Comparison of real data and model predictions for selected countries. The model is able to capture the trend and forecast the future cases. (c-e) sensitivity analysis of model fitting parameters on the output. Increasing *β* decreases the number of cases whereas an increase in *α* an *γ* result in an increase in the number of infected cases.

Using the proposed mathematical model (Equation 1) we modelled the progression of COVID-19 with the data of 46 countries provided by the EU Open Data Portal ^[20]^. By fitting the model to the data set, we deduced the projected final number of COVID-19 cases ∅^*f*^ and the values of fitting parameters *α, β* and *γ* for each country. The variations in *β* as a function of ∅^*f*^ is presented in Figure 3 (a). It can be identified from Figure 3 (a) that there is a strong inverse relationship between the natural reaction of the human body represent by *β* and the projected number of final cases ∅^*f*^. This indicates that in regions where on average people have a strong immune system, the impact of the COVID-19 outbreak may be relatively less influential. Another noteworthy observation from our model is that *α*, related to the diffusive transmission of virus, is inversely proportional to the ∅^*f*^, as shown in Figure 3 (b). This is counter intuitive as one would expect an increase in the size of the pandemic with *α*. Thus, in countries where there are frequent interactions between infected individuals and the population without protective measures (e.g social distancing and masks), the number of projected cases ∅^*f*^ was low. To explain this behaviour, we have presented Figure 3 (c-d) which depicts the negative correlation between *β* and *γ* (Figure 3 (c)), and positive correlation between *β* and *α* (Figure 3 (d)). Interestingly, we have demonstrated mathematically in Figure 3 (c) that as the strength of the natural immune system of the body increases, the growth of the virus is suppressed. Hence, in spite of these individuals having been exposed to the virus, they may not manifest symptoms as the growth of virus in these individuals is restrained. Figure 3 (d) indicates that in regions where there are a greater number of asymptomatic cases, the diffusive transmission of virus *α* remains high. Individuals with asymptomatic infection are entirely unconscious of carrying the virus and, can interact with society without precautions ^[22]^. As a result, we can expect *α* to increase with *β* (Figure 3 (d)). According to Davies et al. ^[23]^, susceptibility to the virus increases with age. Our mathematical model supports the investigation of Davies et al. ^[23]^ suggesting that it is the natural ability of young individuals body to resist the growth of virus that makes them less susceptible in comparison to old aged individuals. We believe that immunity amongst other factors may also explain the inverse relationship between *α* and the predicted number of cases ∅^*f*^. Thus, in countries with high diffusive transmission and low number projected cases ∅^*f*^ (such as those located in Asia and Africa), social interaction may primarily be within age groups that are less susceptible to the virus ^[23]^. This may affect the total number of symptomatic cases recorded since such individuals may be asymptomatic and yet maintain social interactions. However, it is important to note that in these regions the susceptible group of individuals might be at high risk of being infected, as the majority of the population may not modify their behavior. Alternatively, if the outbreak is localized, the number of cases within that zone in relation to the entire population could still be low in spite of the social interactions.

**Figure 3.**
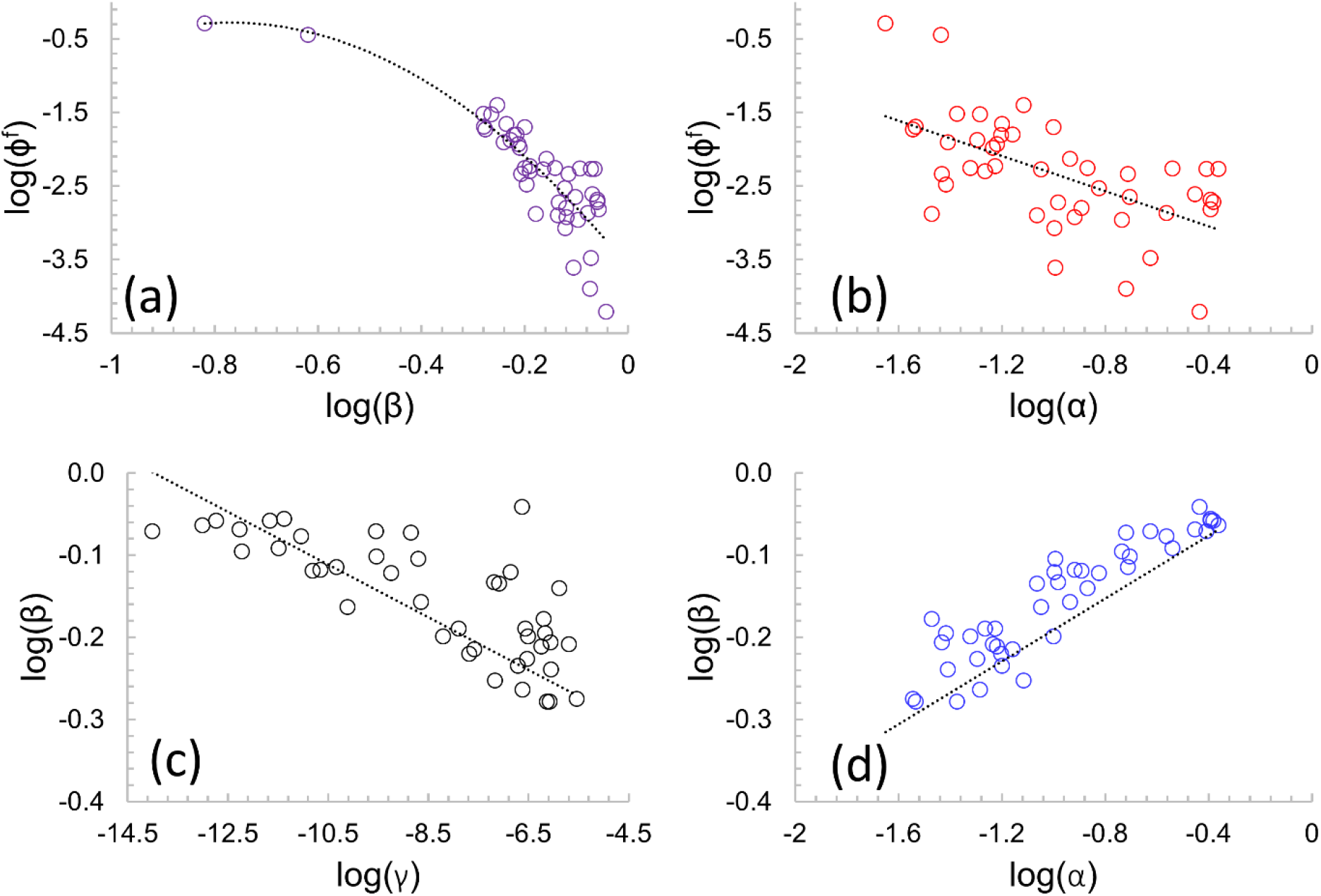
Relationship between model parameters. (a) The negative correlation between natural immunity *β* and the total predicated cases ∅^*f*^, indicating in countries where on average individuals have a strong immune system, the size of the epidemic is less extensive. (b) Decrease in ∅^*f*^ as *α* increases, highlighting an increase in the social interaction of asymptomatic individuals. (c) The *β* is inversely proportional to the *γ*, indicating that as the strength of natural immunity enhances the growth of virus is suppressed. Hence, in these individuals, the virus is not capable of deteriorating their health. (d) Reflects the positive relationship between *β* and *α*. In regions where there is high diffusion transmission of virus individuals many remain asymptomatic and therefore unintentionally transmit the virus.

We have presented an infectious modelling approach for forecasting the course of COVID-19 using fundamental principles of fluid transport. In this physics based model, the infected individual is considered analogous to a fluid containing special species such as ions which spread in the direction of fluid flow, such that the transmission from person to person is dependent on the activity of virus in the host and their immune response. The proposed model demonstrates the relationship between the diffusive transmission of virus, the growth of virus inside human body and the natural immune system of the individual that are the primary factors in controlling the size of the pandemic. Our model suggests that the health/immune response of individuals might be one of the keys to limiting the effects of the pandemic. Despite the simplicity of our approach and the limited data used, the results are promising. In future, other external information such as age demographics, socioeconomics and environmental data will be incorporated to boost the efficacy of the model.

### 2.1 Materials and Methods

Ignoring the advective flux the traditional fluid transport equation is written as ^[19]^; 

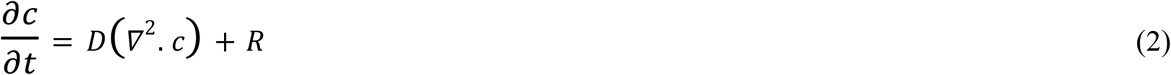

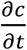 relates to the rate of change in the concentration of species *c* in fluid, *D* is the diffusive flux of species, ▽^2^ is the Laplace indicating spatial coordinates and *R* is the net growth of species. Following our intuition that a fluid is a carrier of species such as salts, an infected individual could also be conceptualized as the carrier of virus. Utilizing this approach we substitute c with ∅ which represents the ratio of total infected cases *I*, over the initial number of susceptible people *S*^*o*^ (assumed as the total population of the specific country), and *R* as 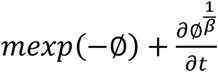.

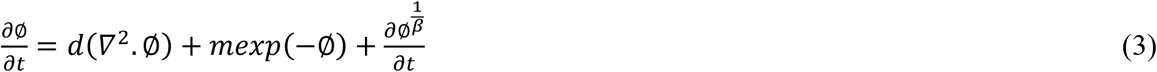

In the above equation, *d* represents the flux of infected individuals, *mexp*(−∅) is the growth of virus within an infected individual, and 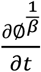 suggests temporal changes in the virus activity due to the response of the natural immune system of individuals. Discretizing Equation 4 using FTCS (Forward Time Centered Space) method results in; 

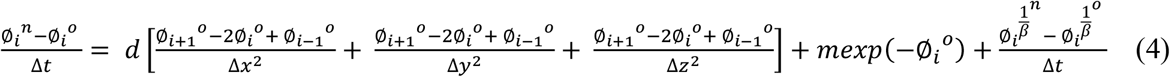

We further simplify Equation 4 by equating ∅_*i*+1_^*o*^ = 0, ∅_*i*−1_^*o*^ = 0 and 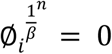. In order to nondimensionalize Equation 4, we substitute 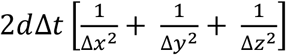 and *m*Δ*t* with *α* and *γ* respectively. This results in the derivation of final equational form, which relates the increase in the infected cases ∅_*i*_^*n*^ − ∅^*o*^ with the transmission of the virus due to diffusion, *α*∅_*i*_^*o*^, the growth of the virus within an infected individual, 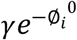, and the suppression of virus growth inside the infected individual due to the natural immune system, 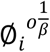. 

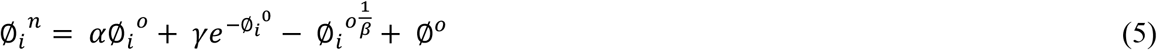

## Data Availability

The data presented in this manuscript will be available freely via sending a request to the corresponding author.

## 3 Acknowledgements

Dr. Harris Sajjad Rabbani would like to acknowledge Dr. Nida Jaleel for the insightful discussion on the topic.

## 6 Author Contributions

H.S Rabbani designed the research and derived the mathematical model. All authors participated in performing the analysis and writing the manuscript.

